# Comparison of Bayesian approaches for developing prediction models in rare disease: application to the identification of patients with Maturity-Onset Diabetes of the Young

**DOI:** 10.1101/2024.01.22.24301429

**Authors:** Pedro Cardoso, Timothy J. McDonald, Kashyap A. Patel, Ewan R. Pearson, Andrew T. Hattersley, Beverley M. Shields, Trevelyan J. McKinley

**Affiliations:** University of Exeter Medical School. Address: Clinical and Biomedical Sciences, RILD Building, Royal Devon & Exeter Hospital, Barrack Road, Exeter EX2 5DW, UK; University of Dundee. Address: Division of Population Health & Genomics, Ninewells Hospital and Medical School, University of Dundee, Dundee DD1 9SY, UK

**Keywords:** MODY, Bayesian modelling, rare diseases, prior elicitation, recalibration

## Abstract

**Background:** Clinical prediction models can help identify high-risk patients and facilitate timely interventions. However, developing such models for rare diseases presents challenges due to the scarcity of affected patients for developing and calibrating models. Methods that pool information from multiple sources can help with these challenges.

**Methods:** We compared three approaches for developing clinical prediction models for population-screening based on an example of discriminating a rare form of diabetes (Maturity-Onset Diabetes of the Young - MODY) in insulin-treated patients from the more common Type 1 diabetes (T1D). Two datasets were used: a case-control dataset (278 T1D, 177 MODY) and a population-representative dataset (1418 patients, 96 MODY tested with biomarker testing, 7 MODY positive). To build a population-level prediction model, we compared three methods for recalibrating models developed in case-control data. These were prevalence adjustment (“offset”), shrinkage recalibration in the population-level dataset (“recalibration”), and a refitting of the model to the population-level dataset (“re-estimation”). We then developed a Bayesian hierarchical mixture model combining shrinkage recalibration with additional informative biomarker information only available in the population-representative dataset. We developed prior information from the literature and other data sources to deal with missing biomarker and outcome information and to ensure the clinical validity of predictions for certain biomarker combinations.

**Results:** The offset, re-estimation, and recalibration methods showed good calibration in the population-representative dataset. The offset and recalibration methods displayed the lowest predictive uncertainty due to borrowing information from the fitted case-control model. We demonstrate the potential of a mixture model for incorporating informative biomarkers, which significantly enhanced the model’s predictive accuracy, reduced uncertainty, and showed higher stability in all ranges of predictive outcome probabilities.

**Conclusion:** We have compared several approaches that could be used to develop prediction models for rare diseases. Our findings highlight the recalibration mixture model as the optimal strategy if a population-level dataset is available. This approach offers the flexibility to incorporate additional predictors and informed prior probabilities, contributing to enhanced prediction accuracy for rare diseases. It also allows predictions without these additional tests, providing additional information on whether a patient should undergo further biomarker testing before genetic testing.

## 1. Background

Clinical prediction models can be useful in rare diseases to aid earlier diagnosis and more appropriate management. However, developing these models can be challenging as suitable data sources for model development may be difficult to acquire. The prevalence of a rare disease in a population-of-interest can be informed by population cohorts [1], but low numbers of cases in these datasets limit the ability to identify risk factors and produce robust predictive models for disease risk in the general population [2]. Case-control studies [3] enrich the study population with more disease cases than a random sample from the population, facilitating more robust estimates of associations between patient features and disease risk using measures such as odds ratios. Furthermore, the rise of rare disease registries [4] makes recruiting larger case numbers for these studies easier. However, from a clinical perspective, disease risk probabilities are more natural metrics than odds ratios for diagnosis or screening purposes, but estimated risk probabilities from case-control data will be overestimated as they are not based on random samples from the general population [5, 6]. A key challenge is, therefore, how to produce well-calibrated estimates of individual disease risk probabilities for rare diseases in the general population, utilising information from different data sources.

Various methods have been developed that borrow information from one population and recalibrate their outputs to be valid in another population [7, 8, 9, 10, 11, 12, 13]. These approaches include simple methods such as adjustments of the likelihood ratio based on the sensitivity and specificity of the test at various thresholds [14] or offset updating to adjust the overall model probabilities according to a more appropriate population prevalence [10, 11]. However, these approaches are limited and would not account for differences in patient characteristics that may occur in different datasets, which could be a particular problem in case-control studies when enriching for a particular disease or when only collecting specific controls, which would ignore more “grey-area” patients that may be seen in a population setting. More complex techniques are available, such as shrinkage methods to adjust the intercept and model coefficients [7, 12], or previous studies could be used to inform the prior belief of model parameters in Bayesian modelling [15]. Although more sophisticated, these approaches would need data from multiple sources that may not be available for rare diseases. In addition, datasets may not always contain the same information for rare diseases, and specific testing or features may only be available to a subset of patients. More flexible approaches are needed that would allow modelling in these situations.

We use a specific motivating example of developing a prediction model for a rare form of diabetes called Maturity-Onset Diabetes of the Young (MODY) that can be used to inform referral decisions for genetic screening for the condition. In this study, we 1) evaluate a range of approaches for appropriately recalibrating model probabilities in prediction models for rare diseases utilising different data sources (including case-control data, prevalence estimates, and population datasets) and 2) develop a Bayesian hierarchical mixture modelling approach which can combine a clinical features risk model with additional informative biomarker test information, utilising prior information from other data sources to account for missing data and ensure that the recalibrated probabilities are clinically plausible given specific test results. This latter approach also allows for predictions for new individuals who do not have biomarker test results (since these are not currently routinely collected for MODY), which greatly facilitates using such a prediction model in clinical practice. We also show how the model can help inform on the utility of additional biomarker testing before making a final screening decision for MODY.

### Motivating example

Our motivating disease system in this manuscript is a rare young-onset genetic form of diabetes called Maturity-Onset Diabetes of the Young (MODY) [16], which is estimated to account for 1–2% of all diabetes cases [17, 18]. MODY is challenging to identify and is estimated to be misdiagnosed in up to 77% of cases [19]. Diagnostic genetic testing is expensive; however, it is crucial to properly diagnose as these patients do not require treatment with insulin injections [20], unlike the most common young-onset form of diabetes, type 1 diabetes (T1D).

Statistical models that use patient characteristics to predict the probability of having MODY can aid decisions regarding which patients to refer for diagnostic MODY testing. One such set of models is routinely used in clinical practice via an online calculator [14] (found at: https://www.diabetesgenes.org/exeter-diabetes-app/ModyCalculator) and has been shown to improve positive test rates of new MODY cases [19]. These prediction models for MODY were developed using case-control data and recalibrated to population prevalences using conversion tables derived from the sensitivities and specificities at different probability thresholds [14]. There are several consequences of this approach for prevalence adjustment: i) the recalibrated probabilities end up being grouped; ii) individuals cannot have a recalibrated probability that is lower than the estimated prevalence in the general population; and iii) the recalibrated probabilities can be sensitive to the choice of grouping used. Addressing these limitations would be important, but the most appropriate approach for adjusting for the prevalence is unclear.

In addition, since the original model development, biomarker screening tests (C-peptide and islet autoantibodies [21, 22]) have become routinely available clinically, and the results of these tests could significantly alter the probability of MODY. C-peptide is a measure of endogenous insulin secretion, and islet autoantibodies are markers of the autoimmune process in T1D. MODY is characterised by non-insulin dependence, so these patients produce significant amounts of their own endogenous insulin (have positive C-peptide), and they do not have the autoimmune process associated with T1D (negative islet autoantibodies), whereas being C-peptide negative (i.e. insulin deficient) or having positive islet autoantibodies is characteristic of T1D. Finding approaches to build these test results into the recalibration would have considerable advantages.

## 2. Methods

### Setting

The diagnosis of MODY requires expensive genetic testing. Currently, patients are referred for diagnostic genetic testing on an ad-hoc basis when the clinician considers a MODY diagnosis. In line with guidelines (ISPAD [23] and NHS genomic testing criteria [24]), criteria for referring can include:

‐ Clinical presentation and patient features (including age at diagnosis, BMI, treatment, measures of glucose control (HbA1c) and family history of diabetes).

‐ Results of biomarker testing (C-peptide and islet autoantibodies)

‐ The use of prediction models in the form of the MODY calculator (can be found at: https://www.diabetesgenes.org/exeter-diabetes-app/ModyCalculator)

### Study population

For model development and recalibration, we used data from two sources comprising patients with confirmed MODY and insulin-treated patients with T1D, the predominant alternative diagnosis in young-onset patients:

### Case-control dataset (Fig. 1a)

This dataset was used to develop the original MODY prediction model [14]. All participants were diagnosed with diabetes between the ages of 1 and 35. T1D was defined as occurring in patients treated with insulin within 6 months of diagnosis [14]. The dataset includes 278 patients with T1D and 177 probands with a genetic diagnosis of MODY obtained from referrals to the Molecular Genetics Laboratory at the Royal Devon and Exeter NHS Foundation Trust, UK. The dataset comprises the following variables: sex, age-at-diagnosis, age-at-recruitment, BMI, parents affected with diabetes and HbA_1c_ (%). No biomarker data are available.

**Fig. 1:**
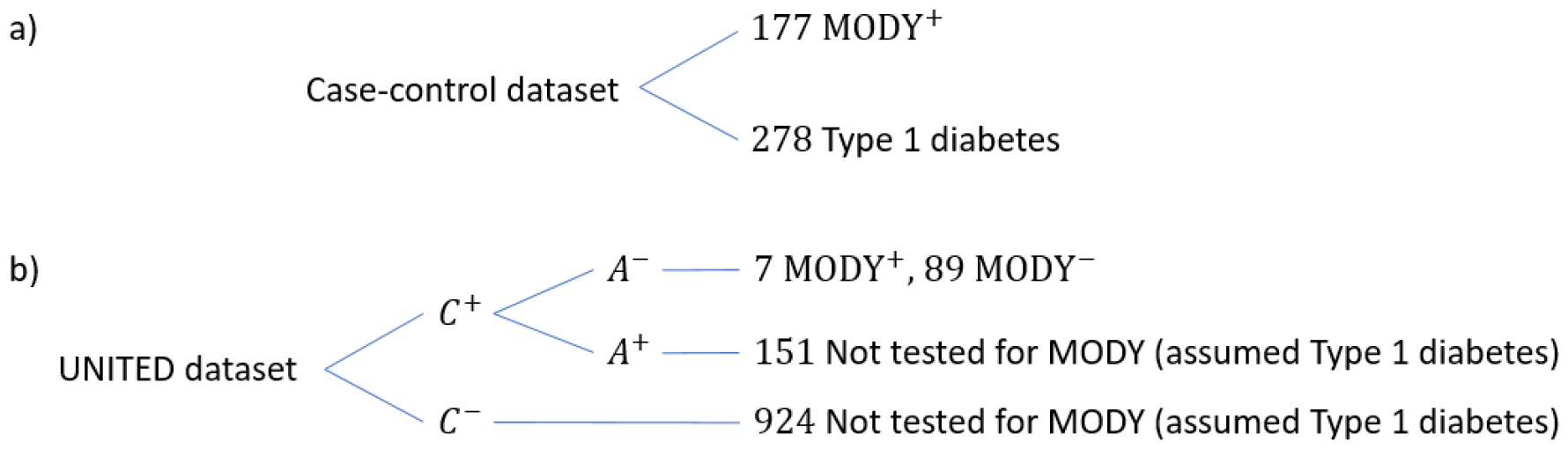
Structure of a) case-control and b) UNITED (population) datasets. MODY^+^ corresponds to a positive test when genetically tested for MODY and MODY^−^ corresponds to a negative test when genetically tested for MODY. C^+^ = C-peptide positive, C^−-^ = C-peptide negative, A^+^ = Antibody positive, A^−^ = Antibody negative

### Population-representative dataset (UNITED – Fig. 1b)

The UNITED study [25] was a population-representative cohort that recruited 62% of all patients with diabetes diagnosed between the ages of 1 and 30 in two regions of the UK (Exeter and Tayside) (n=1418). Due to the expense of genetic testing, a screening strategy with C-peptide and islet autoantibody testing was used to narrow down the cohort eligible for MODY testing (Fig. 1b).

For this model, consistent with the original model [14], we analysed all patients insulin-treated within 6 months of diagnosis, corresponding to 1171 patients, of which 96 were tested for MODY (given that they were C-peptide positive and antibody negative) and 7 MODY cases were diagnosed (Fig. 1b). The dataset is comprised of the following variables: sex, age-at-diagnosis, age-at-recruitment, BMI, parents affected with diabetes and HbA_1c_ (%), with additional C-peptide and islet autoantibodies test results.

### Approaches for recalibration

The analysis in this paper was split into three different scenarios to enable population-appropriate probabilities to be calculated with and without the additional biomarker information:

Scenario a) Clinical features model ignoring biomarker information. For this analysis, we used all patients in the population-representative dataset (UNITED). This scenario assumes all those not MODY tested are in the population cohort, i.e. 7 MODY positive patients and 1,164 MODY negative (of which 1,075 were not tested for MODY but are assumed to be MODY negative for the analysis since the biomarker results are inconsistent with MODY).

Scenario b) Clinical features model in only those pre-screened to be at increased probability of MODY based on the biomarkers. This included 96 patients, of which 7 are MODY positive and 89 MODY negative. This scenario only analyses patients in the population cohort who had genetic testing (i.e. tested C-peptide positive and autoantibody negative), so it provides more appropriate model probabilities in patients with these test results indicating a higher risk of MODY, but simply rules out MODY (does not provide a probability) in those who are C-peptide negative or antibody positive.

Scenario c) Model fully incorporating both clinical features and biomarker information. We analysed all patients in the population cohort and included biomarker information. For this analysis, we included 96 patients who had testing for MODY (7 MODY positive and 89 MODY negative) and 1,075 patients who did not have testing for MODY. The biomarker information of those not MODY tested was used to more appropriately adjust the model probabilities (151 C-peptide positive and autoantibody positive, 924 C-peptide negative) (Figure 1b).

We explored six approaches for producing predictions using different degrees of data availability, which fall into three groups:

1. Approaches that only utilise case-control data and adjust to a known population prevalence: *Original* and *Offset*.
2. Approaches that utilise a case-control dataset and additional calibration dataset (e.g. the UNITED population dataset in this study): *Re-estimation* and *Recalibration*.
3. Approaches that utilise additional data on informative diagnostic tests and provide biologically plausible constraints: *mixture model approaches* (for both *Re-estimation* and *Recalibration*). This mixture model splits individuals into two groups according to their diagnostic test information (a C-peptide negative or antibody positive group: ; and a C-peptide positive *and* antibody negative group:). We use an informative prior distribution to constrain the probability of having MODY in the group and use one of the other recalibration methods in the group.

The Supplementary Materials Notation section contains a glossary of mathematical symbols used throughout the article. We fit these models using the package **NIMBLE** [26, 27] (version 1.0.1) in the software R [28] (version 4.3.2).

### 1. Training dataset only approaches

#### Training data model

Let 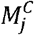 be a binary variable denoting whether an individual *j* in the case-control data set has MODY or not, such that

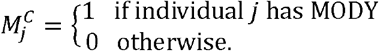

We then model:

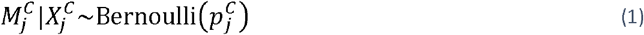

where the log odds are given by:

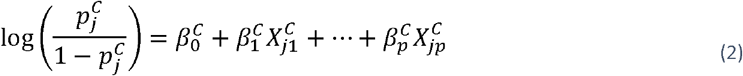

with 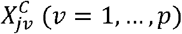 a set of *p* covariates for individual *j*. We put independent vague Normal (*µ* = 0, sd = 10) priors on the regression parameters.

The posterior for this model then takes the form:

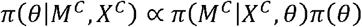

where 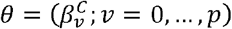, with *π*(·) denoting the relevant probability (density) mass functions derived above for the model and joint prior distribution.

#### Original approach

This method was implemented by Shields *et al*. (2012) [14] during the development of the original MODY prediction model. The approach fitted a model to a case-control dataset using the patients’ characteristics and used the relationship:

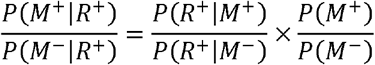

or

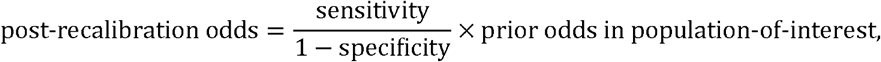

where *M* ^+^ is the event that the patient has MODY, and *R*^+^ is whether a hypothetical “test” is positive (and similarly for *M*^−^ and *R* ^−^). In this case, *R*^+^ is derived by applying a threshold, *P*^*^, to the predicted probabilities 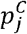 obtained from a training model (see equation (*2*)) for a given individual *j*, such that an individual is classed as positive if 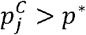 and negative otherwise.

Therefore, for a given choice of *P*^*^, estimates of the sensitivity *P* (*R* ^+^ | *M* ^+^), and specificity, *P* (*R* ^−^ | *M* ^−^), of these classifications at a range of thresholds were calculated using the case-control data. *P* (*M* ^+^) is then chosen as an estimate of the prevalence of MODY in the general population, which in the original model was given by 0.7% [14], which assumed no knowledge of biomarker test results. In this paper, we adjusted slightly differently depending on scenario a) or scenario b). In scenario a), we estimated the pre-test probability to be 0.6% (informed by the prevalence of MODY in the UNITED dataset). For scenario b), we estimated the pre-test probability to be 7.3% (informed by the prevalence of MODY in those who were C-peptide positive and antibody negative in UNITED).

For a new individual in the general population, with covariates 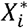 say, then one can derive an estimate for 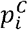 (based on equation (*2*)) as

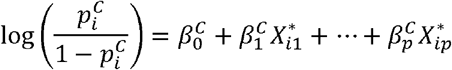

before using Table 1 to map their predicted 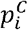 from the case-control model to a *recalibrated probability* of having MODY in the general population.

**Table 1:** Probability conversion table for MODY using the Original method with an adjusted pre-test probability. (A) Model using clinical features only adjusting to population prevalence of MODY (0.6%), (B) model using clinical features but adjusting to population prevalence of MODY based on patients who are C-peptide positive and antibody negative (7.3%). Parentheses are used to signify that an endpoint value is not included. Bracket are used to signify that an endpoint value is included.

| (A) Original model adjusting probabilities for prevalence [scenario a)] (pre-test probability = 0.6%) |  | (B) Original model adjusting probabilities assuming patients have biomarker tests suggesting increased risk of MODY [scenario b)] (pre-test probability=7.3%) |  |
| --- | --- | --- | --- |
| Case-control model probability (%) | Post-recalibration probability (%) | Case-control model probability (%) | Post-recalibration probability (%) |
| (0, 10) | 0.6 | (0, 10) | 7.3 |
| [10, 20) | 1.6 | [10, 20) | 17.9 |
| [20, 30) | 2.3 | [20, 30) | 23.2 |
| [30, 40) | 3.4 | [30, 40) | 31.6 |
| [40, 50) | 4.2 | [40, 50) | 36.6 |
| [50, 60) | 5.5 | [50, 60) | 43.1 |
| [60, 70) | 6.2 | [60, 70) | 46.5 |
| [70, 80) | 7.1 | [70, 80) | 49.9 |
| [80, 90) | 10.9 | [80, 90) | 61.6 |
| [90, 100] | 45.4 | [90, 100] | 91.6 |

#### Albert Offset approach

This approach was proposed by Albert (1982) [10] and similarly to the method above, leverages the relationship:

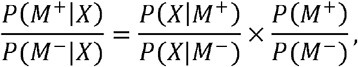

where *X* is a set of explanatory variables. In words:

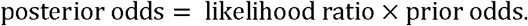

For the training data (C – case-control dataset), we use the same model for 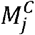 and 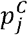 as before (see equations (*1*) and (*2*)), and then the idea is that if we know the *disease odds* in the *training data*, then we can re-write equation (*2*) as:

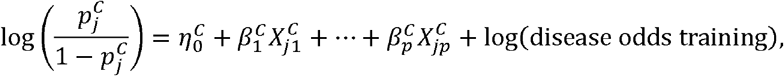

hence the original 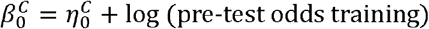. Therefore, under the assumption that *likelihood ratio for any set of covariates is the same* in the training and calibration datasets, then for a new individual *i* in the general population, with covariates 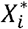, we can recalibrate as:

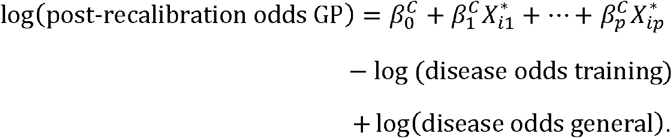

This approach gives individual-level recalibration probabilities that do not rely on thresholding. The *Albert Offset* approach requires a training dataset for fitting the original model and an estimate of the *disease odds* in both the training data and the population-of-interest. For this cohort, as before, you could adapt the offset based on the prevalence of MODY of 0.6% based on scenario a) or 7.3% based on scenario b). We also explore an example where the likelihood ratio assumption is not maintained between datasets for illustrative purposes. We put independent vague Normal (*µ* = 0) priors on the regression parameters.

### 2. Population-representative dataset approaches

#### Re-estimation approach

This approach fits a new model directly to the population-representative dataset (UNITED), ignoring the case-control dataset entirely. Given sufficient cases and controls in a given dataset, this model fitted using, e.g. maximum likelihood, will give asymptotically unbiased estimates for the odds ratios and probabilities. However, for rare diseases, one would have to have very large sample sizes to get sufficient numbers of cases to develop an entirely new model. As a comparison, we use the model structure developed in the case-control dataset and then refit the model to the population-representative dataset (UNITED). Here, we denote the MODY status for individual *i* in the UNITED structure developed in the case-control dataset and then refit the model to the population dataset as 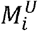, and model this as

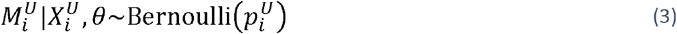

where

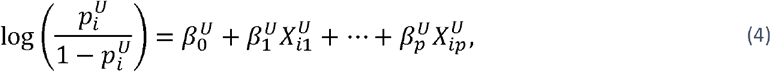

with 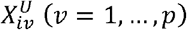 a set of *P* covariates for individual *i*. We place independent vague Normal (*µ* = 0, sd = 10) priors on the regression parameters.

The posterior for this model then takes the form:

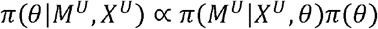

where 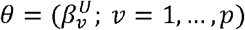, with *π*(·) denoting the relevant probability (density) mass functions derived above for the model and joint prior distribution.

#### Recalibration approach

In the context of the models developed here, the *Recalibration* approach [*7*] uses a model fitted to the case-control dataset to generate predictions of the linear predictor for each individual in the population-representative data set (UNITED). In the data, for individual *j*, 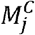 and 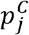 are modelled as before (see equations (*1*) and (*2*)), and then for each individual *i* in the *calibration* dataset (UNITED), with predictors 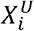, the linear predictors 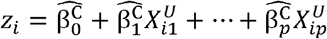 are calculated, where 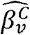 is a point estimate of the *v*^th^ regression parameter from the case-control model. These *z*_*i*_ terms are then used as covariates in a second (shrinkage) model:

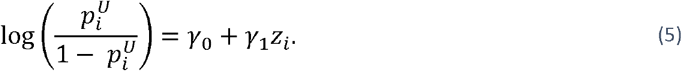

This approach [7] can have the effect of scaling the odds ratios and intercept terms where necessary, and a side-effect is that if no recalibration is required, then *γ*_0_ = 0 and *γ*_1_ = 1. Again, these approaches could be built using scenarios a) and b), dependent on the assumptions we are willing to make with UNITED. The method used by Steyerberg et al. (2004) [7] uses the point predictions for *z*_*i*_ based on the maximum likelihood estimates from the case-control data, which ignores the uncertainty in the estimations of *z*_*i*_. Instead, we develop a joint Bayesian hierarchical model where we simultaneously fit both models and propagate the uncertainties directly from the case-control model to the recalibration model [29]. We put independent vague Normal (*µ* = 0, sd = 10) prior distributions on the regression parameters, with a Normal (*µ* = 0, sd = 10) prior for *γ*_0_ and a Normal (*µ* = 1, sd = 10) prior for *γ*_1_.

The posterior for this joint model then takes the form:

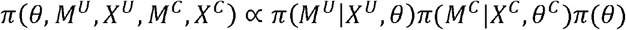

where *θ* = (*θ*^U^, *θ*^C^) corresponds to the full vector of parameters, with *θ*^U^ = (*γ*_0_, *γ*_1_) and 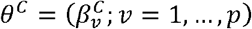, with *π* (·) denoting the relevant probability (density) mass functions derived above for the different component models and joint prior distribution.

### 3. Mixture model approach

One area of development in this manuscript is how to incorporate biomarker test information into the model when the biomarker tests can place very strong constraints on the post-recalibration probabilities depending on their specific values. For example, a simple way to include a binary test result would be to add another covariate into the linear predictor in one of the previous methods. In the analysis, the biomarker data only exists in the calibration data (UNITED) but not the training data (case-control), so this approach would only use information from the calibration data to estimate the parameters relating to the biomarkers. Since there are few cases in the calibration data, this would necessarily result in large standard errors for the estimated effects and could lead to biologically implausible estimates. For example, an individual who is C-peptide negative *or* antibody positive can be considered to have a very low probability of having MODY, justified through prior data and biological plausibility (C-peptide negativity means that an individual is producing negligible amounts of their own insulin, which defines T1D). In clinical practice, an individual with these biomarker results would be treated as having T1D, which is equivalent to assuming that the probability of having MODY given these results is exactly zero. However, this approach does not allow for the rare (but possible) event that an individual has a positive genetic MODY test but is antibody positive or C-peptide negative (which would ideally also allow for imperfect sensitivities and specificities of the biomarker tests).

Using the mixture model approach in scenario c), it is possible to incorporate a non-zero prior probability of having MODY in these cases, where we use independent data sets to inform the prior distribution for this probability. We note that the mixture model allows for different prior constraints to be used for different subsets of the data: here the prior probability of having MODY is very low for C-peptide negative or antibody positive individuals [21, 22], but is not similarly constrained for C-peptide positive and antibody negative individuals. Similar ideas could be used for other diseases where the prior information may not be as strong.

For the UNITED data, we let

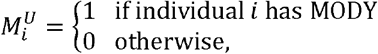

with

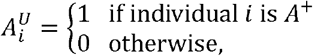

and

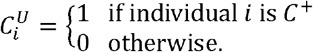

We then set:

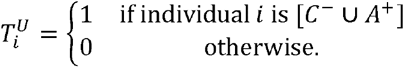

Letting 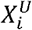 be a vector of additional covariates for individual *i*, we can model 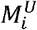 as

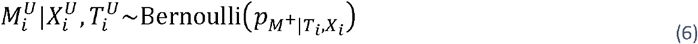

where

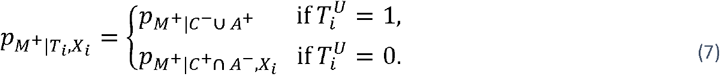

We model 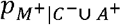 using a Beta (*α* = 2.2, *β* = 7361.3) prior probability distribution (see Supplementary Materials Prior elicitation section for a justification of this choice). We then model 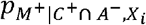 differently, depending on whether we use the *Re-estimation* or *Recalibration* approaches (see below).

#### Re-estimation approach

For the *Re-estimation* approach we model

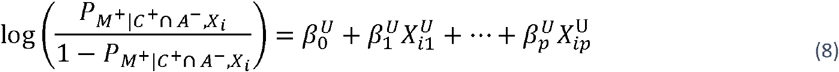

and to finalise, we put independent vague Normal (*µ* = 0, sd = 10) priors on the regression parameters.

#### Recalibration approach

For the *Recalibration* approach we also utilise the case-control data. If we let 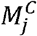 be the MODY status for individual *j* in the case-control dataset, with vector of covariates 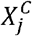, then 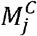 and 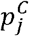 are modelled as before (see equations (*1*) and (*2*)). Then, for individual *i* in the UNITED dataset (with 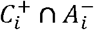), we model

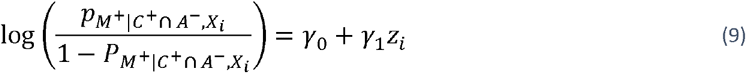

where

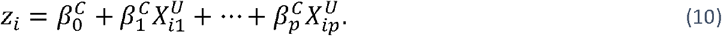

#### Incorporating biomarker test results

To allow for predictions in the absence of biomarker test results (which are not routinely collected in clinical practice), we model

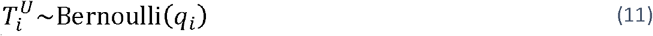

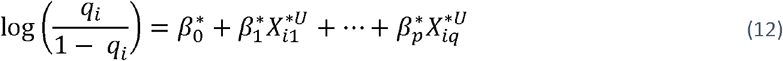

with 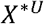 comprised of the variables BMI, age-of-diagnosis, age-of-recruitment and parents affected with diabetes (here we use restricted cubic splines with 3 knots to model the continuous variables). In this case the predicted probability of MODY for an individual with unknown test results will be a weighted average of the 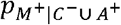 and 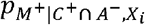, weighted by the probability of being *C*^−^ ⋃ *A*^+^ based on suitable individual-level characteristics. We place independent vague Normal (*µ* = 0, sd = 10) prior distributions on the regression parameters, with a Normal (*µ* = 1, sd = 10) prior on *γ*_0_ and a Normal (*µ* = 1, sd = 10) prior on *γ*_1_.

For the *Re-estimation mixture*, the posterior then takes the form:

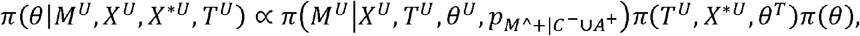

where 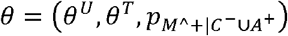 corresponds to the full vector of parameters, with 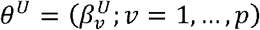 and 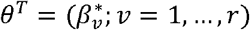 with π(·) denoting the relevant probability (density) mass functions derived above for the different component models and joint prior distribution.

For the *Recalibration mixture*, the posterior then takes the form:

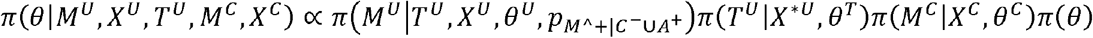

where 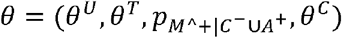 corresponds to the full vector of parameters, with 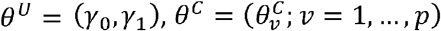 and 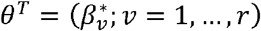, with π(·) denoting the relevant probability (density) mass functions derived above for the different component models and joint prior distributions.

#### Assessment of model performance, calibration and stability analysis

In scenario a), we validate fitted probabilities for all patients in UNITED (setting those with missing MODY testing to MODY^−^). In scenarios b) and c), we only validate fitted probabilities on *C*^−^ ∪ *A*^+^ patients as these were the only patients who had pre-screening based on biomarkers and had genetic testing of MODY genes. The area under the receiver operating characteristic (AUROC) curve was used as a measure of overall discrimination performance. Calibration curves were plotted to visualise how well the predicted probabilities were calibrated against the observed data. For the calibration curves, predicted probabilities were grouped by quintiles and plotted against the observed probability of positive individuals within each quintile. To assess convergence, we monitored the available parameters for evidence of convergence and Gelman-Rubin 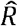 values [30]. Further validation procedures are explained in the Supplementary Materials Stability analysis section.

## 3. Results

### Comparing datasets

In the case-control dataset, 177 out of 455 patients had MODY, leading to an enriched proportion with MODY of 40%. In contrast, in the recalibration population (UNITED) cohort, 7 out of 1171 patients (0.6%) had MODY, much more consistent with the prevalence of MODY in the population. The characteristics of patients in the two datasets were broadly similar (sFig. 1).

### Models and their recalibration from the 6 different approaches

All models in this study converged quickly, so we ran four chains of 500,000 iterations, with the first 300,000 discarded for burn-in in each case (sFig. 2-4).

The first, recalibration approach, the *Original* approach achieved an 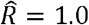 for all parameters. As expected, the choice of prevalence used for recalibration affected the conversion probabilities. Table 1 shows the different *post-recalibration probabilities* of having MODY using the different prevalences for both scenarios a) and b), with post-recalibration probabilities more appropriately higher in scenario b) to allow for the biomarker results in those who were C-peptide positive and antibody negative.

Table 2 describes the model parameter estimates for the *Albert Offset* and *Re-estimation* approaches in scenarios a) and b). The *Albert Offset* approach achieved an 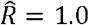, and the *Re-estimation* approaches achieved an 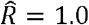 for all parameters. Coefficients were quite different in the various approaches.

**Table 2:**
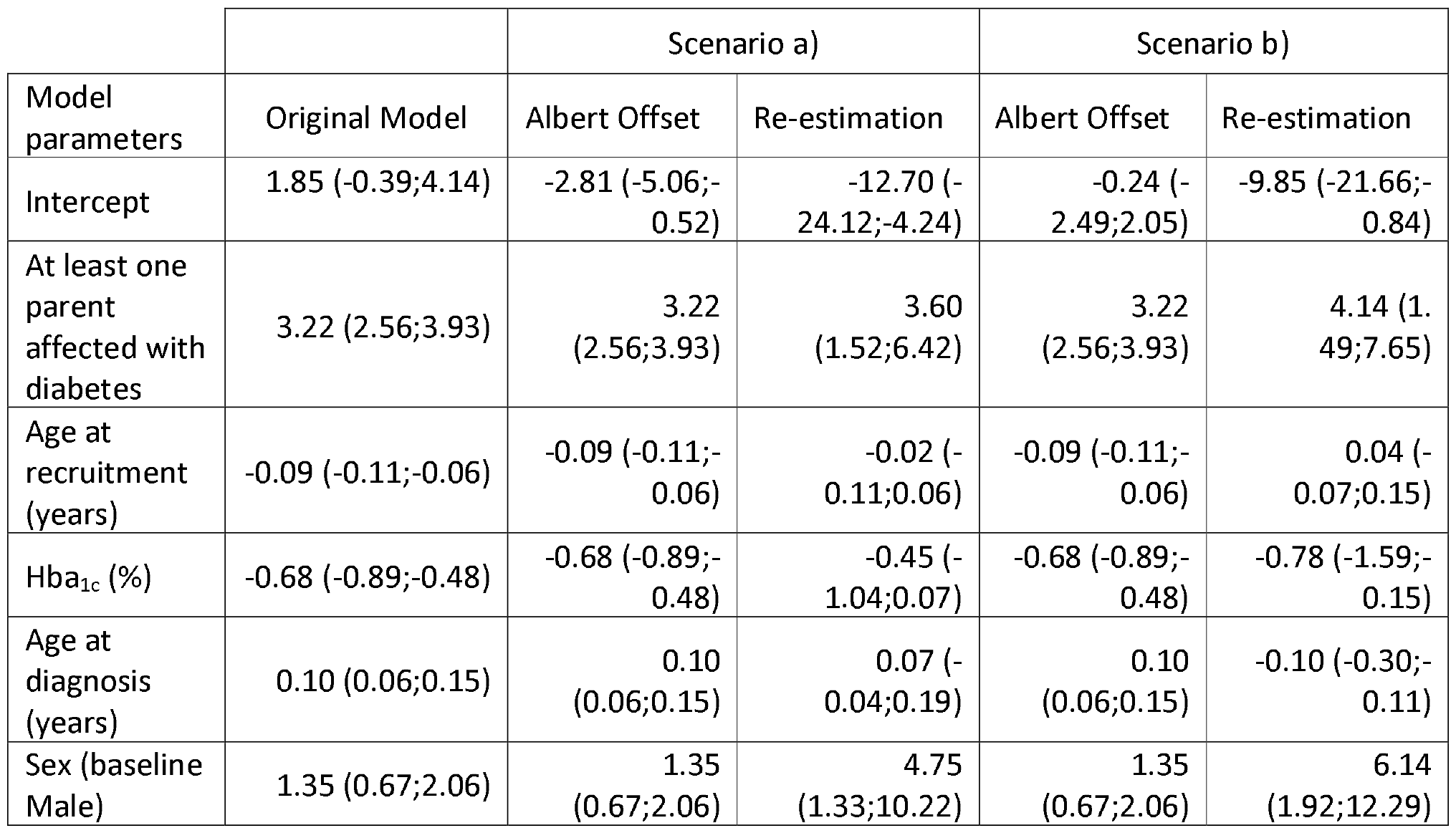
Model parameter estimates for Albert Offset and Re-estimation approaches for scenarios a) and b). Scenario a) adjusts model probabilities based on the population prevalence. Scenario b) adjusts model probabilities for just those who were C-peptide positive and autoantibody negative (n=96). The numbers in the parentheses correspond to the 95% credible intervals.

The *Recalibration* approach achieved an 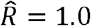 for all parameters. In scenario a), estimates were *γ*_0_ = −4.39 (95%CI -5.41;-3.56) and *γ*_1_ = 0.96 (95%CI 0.49;1.54) and in scenario b), the estimates were *γ*_0_ = −2.26 (95%CI -3.33;-1.37) and *γ*_1_ = 0.86 (95%CI 0.31;1.57).

For scenario c), fully incorporating the biomarker information into the model, probabilities could be obtained using the *Recalibration* and *Re-estimation* mixture approaches. The *Recalibration mixture* approach achieved an 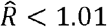 for all parameters, with an estimated *γ*_0_ = −2.26 (95%CI - 3.33;-1.37) and *γ*_1_ = 0.86 (95%CI 0.32;1.58). The *Re-estimation mixture* approach achieved an 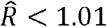. The model that estimates *T* achieved an AUROC of 0.76 (95%CI 0.75;0.77) in both mixture approaches, with model parameters described in sTable 1.

### Discrimination and calibration of the models developed using the 6 different approaches

All approaches led to good model discrimination, with the *Re-estimation* approaches having the highest AUROC (Table 3).

**Table 3:** Area under the receiver operating characteristics (AUROC) for all approaches in both scenarios. In scenario a): ignoring biomarker information (n=1171); Scenario b): only analysing patients which tested C-peptide positive and autoantibody negative (n=96); Scenario c): analyse all patients, adjusting for biomarker results in the model (n=1,171, validated in n=96). CI: credible interval

|  | Approach | Mean | CI: 2.5% | CI: 97.5% |
| --- | --- | --- | --- | --- |
| Scenario a) | Original | 0.93 | 0.90 | 0.94 |
|  | Albert Offset | 0.93 | 0.90 | 0.94 |
|  | Re-estimation | 0.94 | 0.90 | 0.96 |
|  | Recalibration | 0.93 | 0.92 | 0.94 |
| Scenario b) | Original | 0.86 | 0.81 | 0.89 |
|  | Albert Offset | 0.86 | 0.82 | 0.89 |
|  | Re-estimation | 0.92 | 0.89 | 0.94 |
|  | Recalibration | 0.87 | 0.84 | 0.90 |
| Scenario c) | Re-estimation mixture | 0.92 | 0.87 | 0.94 |
|  | Recalibration mixture | 0.87 | 0.84 | 0.90 |

In scenario a), for the approaches that used only the case-control dataset and adjusted for a known prevalence, the *Original* approach overestimated the observed probability of MODY in the UNITED population and had large uncertainty at higher percentages. In contrast, the *Albert Offset* approach slightly underestimated the observed probability of MODY in the UNITED population. Looking at the approaches that used the population-representative dataset (UNITED), both the *Re-estimation* and *Recalibration* approaches slightly underestimated the observed probability of MODY with slightly more uncertainty in the predictions from the *Re-estimation* approach (Fig. 2).

**Fig. 2:**
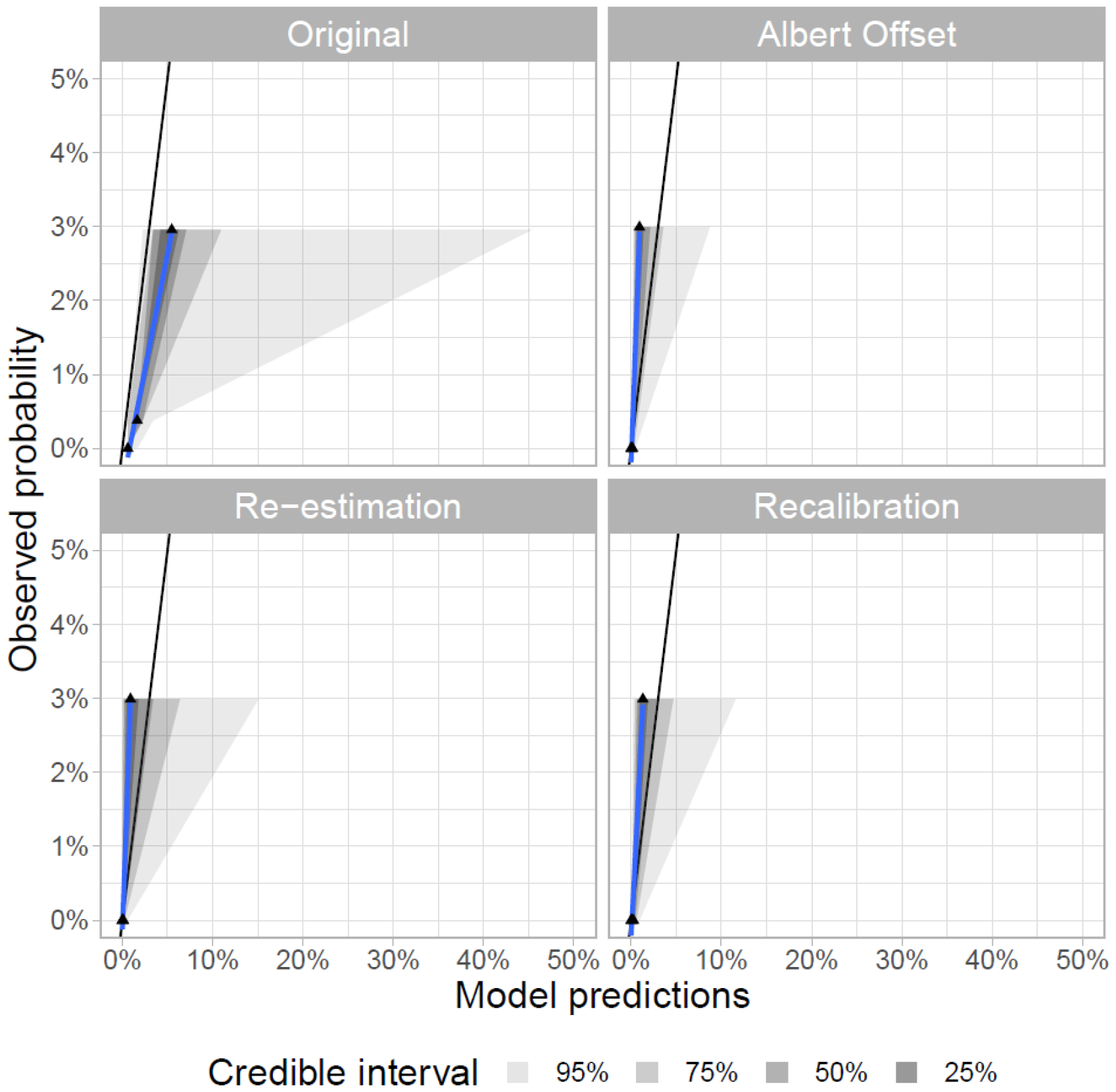
Calibration of scenario a) in UNITED. Scenario a): assume all not MODY tested are MODY^−^ (based on strong clinical knowledge (n=1,171).

In scenario b), for the approaches that used the case-control dataset alone, with adjustment for known prevalence, the *Original* approach overestimated the observed probability of MODY in the UNITED population. In contrast, the *Albert Offset* approach was able to calibrate well. Looking at the approaches that used an additional calibration dataset (UNITED), both the *Re-estimation* and *Recalibration* approaches calibrated well, but the *Re-estimation* approach demonstrated more uncertainty in the probability predictions (Fig. 3). In scenario c), the *Re-estimation* and *Recalibration mixture* approaches demonstrated similar performance to the equivalent models that did not use a *mixture model* approach, with similar levels of uncertainty in probability predictions (Fig. 3). In this case, the *Albert Offset* method worked well, but it relies on the assumption that the likelihood ratio is the same in the two populations. For illustrative purposes, we also provide an example setting where the likelihood ratio is different between the training and calibration datasets (violating the assumption). In this latter example, the *Albert Offset* approach fails to calibrate well. In contrast, the *Recalibration* approach can scale the odds ratios and calibrates well (sFig. 5), so this method would be preferred if a recalibration dataset is available.

**Fig. 3:**
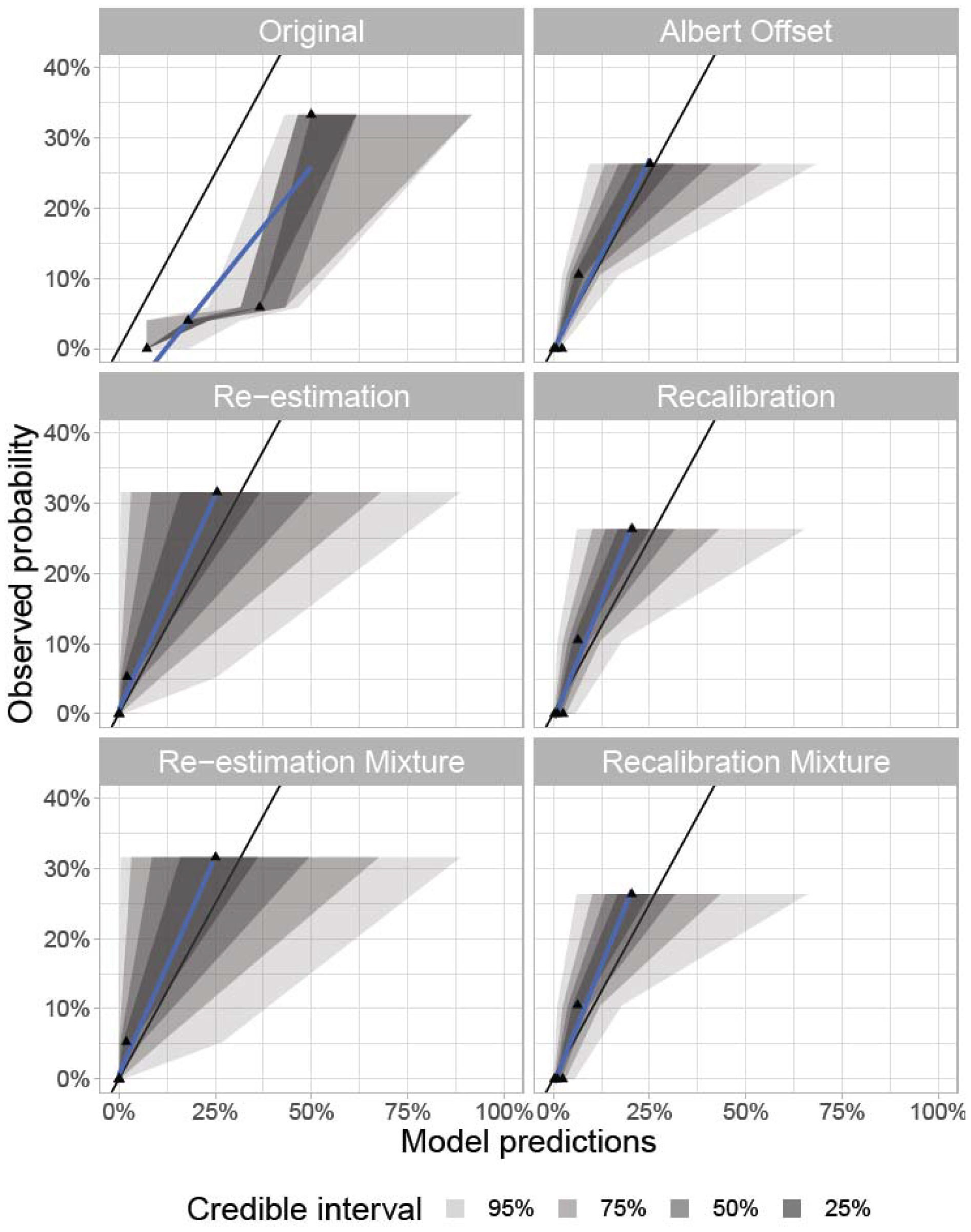
Calibration of scenario b) and c) in UNITED. Scenario b): only analyse patients which tested C-peptide positive and autoantibody negative (n=96) – Original, Albert Offset, Re-estimation and Recalibration approaches. In scenario c): analyse all patients (n=1,171, validated in n=96) – Re-estimation mixture and Recalibration mixture approaches.

### Stability plots for the mixture models in scenario c)

The bootstrap stability test was made for both mixture approaches. Both mixture approaches were ran 50,000 iterations with the first 30,000 discarded for burn-in, with an average 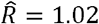 (95% 1.0;2.1) for the Re-estimation mixture approach and an average 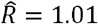 (95% 1.0;1.6) for the *Recalibration mixture* approach (the higher 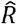 values occurred in bootstrapped datasets with less than 3 positive MODY cases, but this only occurred in 8/1000 datasets and made no difference to the plots in Figure 4, and so we left these runs in). Both recalibration approaches showed some variability in the estimated probabilities, with the *Re-estimation mixture* approach demonstrating higher uncertainty across all estimated probability levels (Fig. 4). However, we can see that because the *Recalibration mixture* approach borrows weight from the case-control data, the estimates were more stable than the *Re-estimation mixture* method. We also noted that by using the hierarchical modelling approach, the *Recalibration mixture* model uncertainty included the uncertainty in the case-control predictions. Thus, these uncertainty estimates are larger than a model where this additional predictive uncertainty is ignored.

**Fig. 4:**
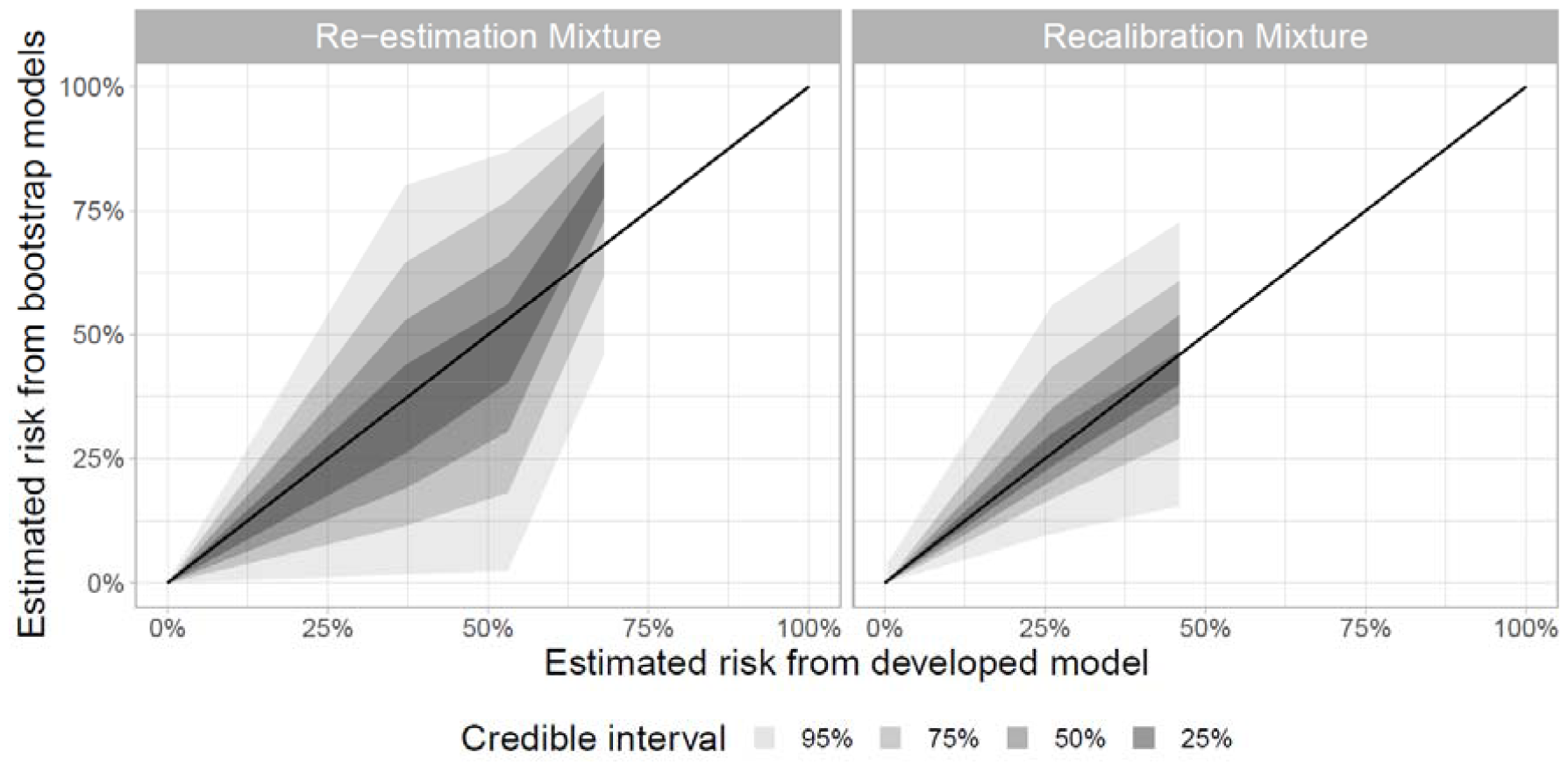
Stability plots for Re-estimation and Recalibration mixture approaches. Estimations of MODY probability from bootstrapped models are plotted against estimated MODY probabilities from the developed model.

### Final recalibrated probabilities

The approach chosen for our final models was the *Recalibration mixture* approach, which incorporated the most information with the lowest uncertainty in probability predictions. The mixture model ensures that those with biomarkers consistent with T1D (the *C*^−^ ∪ *A*^+^ individuals) are predicted to have a very low probability of MODY, consistent with independent prior information. Fig. 5 shows the predicted probability of MODY in the remaining *C*^+^ ∩ *A*^−^ individuals. Considering only 0.6% of the cohort had MODY, the model produced a wide range of probabilities. Most non-MODY cases were predicted to have a low probability of MODY, with 97.2% (1,132/1,164) of individuals having an upper 95% CI probability of MODY under 10%. In contrast, 7 out of the 7 MODY cases had an upper 95% CI probability >10%. This would mean that if using a >10% threshold to initiate MODY testing for the population, 39 patients would be tested, giving a positive predictive value of 17.9% (Fig. 5), equivalent to the *Original* approach.

**Fig. 5:**
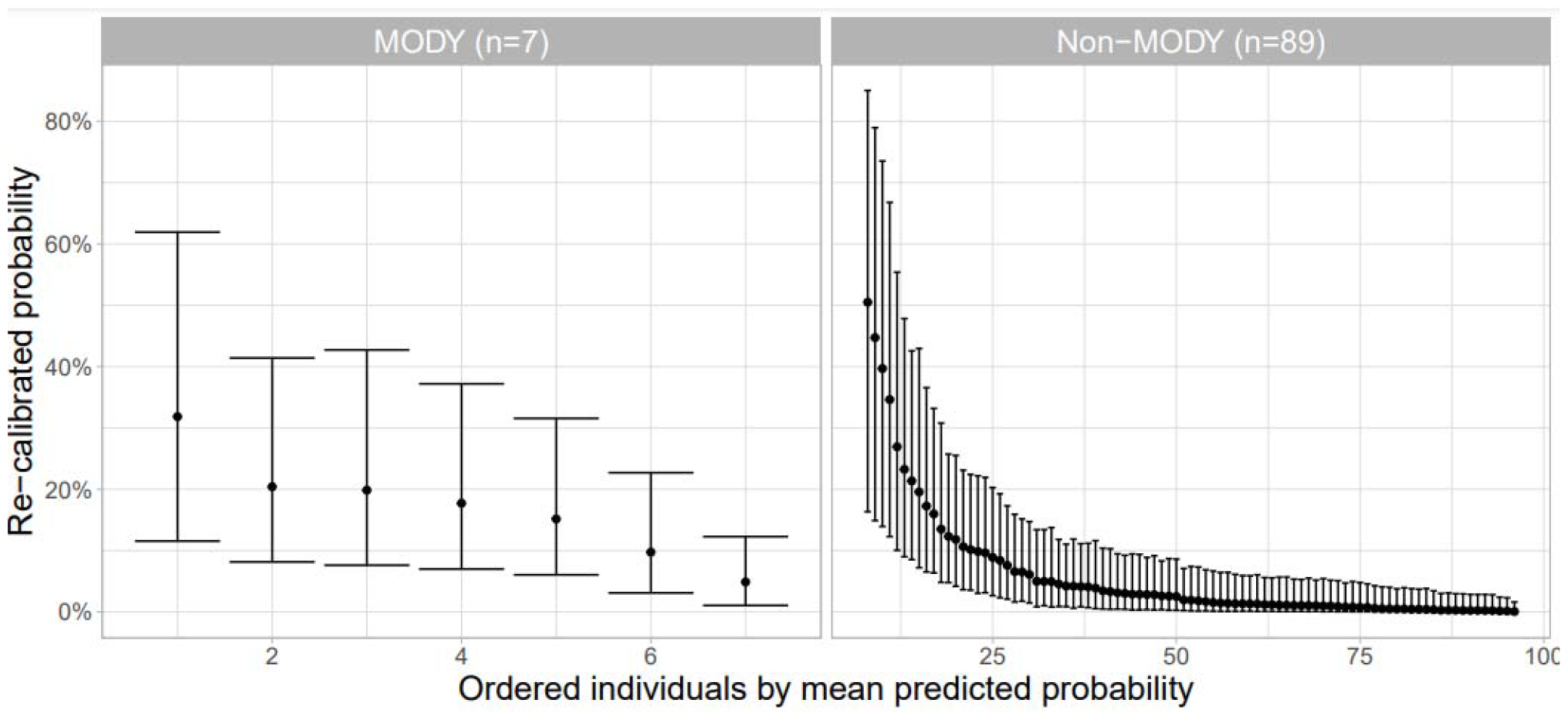
Estimated probabilities of MODY from the Recalibration mixture model in C-peptide positive, antibody negative patients, split by whether patients tested positive for MODY or not. All patients with negative C-peptide or positive antibodies (n=1,075) had probabilities close to 0 and are not shown.

## 4. Discussion

This paper explored recalibration methods for adapting a statistical model from case-control data to the general population for rare disease prediction.

We have shown that the calibration of disease risk probabilities can be improved via various methods, and in particular, our results show the added benefits of utilising a secondary (recalibration) dataset that corresponds to a random sample from the general population despite there being few cases in the latter. In addition, the recalibration data contains additional information on biomarker tests, which are highly informative about disease risk, but only for certain subsets of test results; because of this, some biomarker information is only available for subsets of individuals. Our *Recalibration mixture* model allows the inclusion of (incomplete) biomarker information and informative prior information (derived from previous studies) about disease risk for specific subsets of test results to ensure clinically valid risk probabilities in those cases.

The *Recalibration mixture* model has several other advantages. It allows for predictions to be made in clinical practice even if the biomarker information is not available. We can also propagate parameter uncertainties from the case-control model to the recalibrated population predictions by utilising the Bayesian framework. This gives a more robust estimate of the underlying predictive uncertainty than classical models that ignore this uncertainty. Furthermore, the predictions for individuals without biomarker test information also propagate the uncertainties from the missing information. Finally, since this model is used to help inform which individuals should be screened for MODY using expensive genetic testing, for those individuals who have missing biomarker information, we show how the mixture model can also be used to inform clinicians about the added utility of performing a biomarker test before making a final decision of whether to send individuals for genetic testing. Although highlighted with a specific application, these ideas could be adapted to other rare diseases.

We compared several approaches for recalibrating probabilities when developing prediction models for rare diseases. We showed that the *Original* method tends to overestimate the probabilities in the general population, but that the *Albert Offset* [10], *Re-estimation* and Recalibration [7] approaches achieve good calibration of MODY probability predictions in both the model of the overall population and also in the model examining only the subset who were *C*^+^ ∩ *A*^−^ (those genetically tested for MODY). The *Albert Offset* [10] and *Recalibration* [7] approaches achieved the smallest uncertainty around the observed probability of MODY. The *Recalibration mixture* model showed stability in our study and was the only approach that appropriately constrained the probability of MODY in *C*^−^ ∪ *A*^+^ individuals to be consistent with the strong prior information available in this setting. When developing prediction models for rare diseases in practice, different approaches will be plausible in different scenarios based on the available data sources. Table 4 provides an overview of the advantages and disadvantages of all modelling approaches explored in the manuscript.

**Table 4:** Summary of key characteristics of the several modelling approaches.

| Modelling Approaches | Advantages | Disadvantages |
| --- | --- | --- |
| Original | - Model can be built in one training dataset (e.g. case-control) but probabilities can be updated based on information of prior probability from other studies. | - Individuals cannot have a recalibrated probability lower than the estimated prevalence in the general population. |
|  |  | - Probabilities are aggregated into groups. |
|  |  | - The recalibrated probabilities can be sensitive to the choice of grouping used. |
| Albert Offset | - Model can be built in one training dataset (e.g. case-control) and probabilities can be updated based on information of prior probability from other studies. | - Assumes the likelihood ratio for any given set of covariates is the same in the training data and calibration/target population. |
| Re-estimation | - Model can be built on one training dataset (population-representative) so no recalibration necessary. | - Requires a large sample size from the general population if model development required. |
|  |  | - A lot of uncertainty in a rare disease setting due to very low number of positive cases. |
| Recalibration | - Scales odds ratios allowing for easy recalibration so that the model can be used in different settings (e.g.: general population or lab referral usage). | - Requires two datasets: a training dataset (e.g. case-control) and recalibration dataset (e.g. general population). |
| Re-estimation mixture | - The model can be specified to include additional information on biomarker testing. | - Requires a large sample size from the general population. |
|  | - Model can be built on one training dataset (population-representative) so no recalibration necessary. | - A lot of uncertainty in a rare disease setting due to very low number of positive cases. |
| Recalibration mixture | - The model can be specified to include additional information on biomarker testing. | - Requires two datasets: a training dataset (e.g. case-control) and recalibration dataset (e.g.: general population). |
|  | - Scales odds ratios allowing for easy recalibration so that the model can be used in different settings (e.g.: general population or lab referral usage). |  |

When only a training dataset (case-control dataset in our setting) is available, and the aim is to adjust probabilities based on population prevalence, then the *Albert Offset* approach was the preferred method as it estimated the probability of MODY well in both our scenarios, with reasonable uncertainty in the predictions. In contrast, the *Original* approach [14] relies on thresholding probabilities and overestimates the probability of MODY in both scenarios. The *Albert Offset* approach has also been compared to other recalibration methods in other studies. Chan *et al*. (2008) had similar findings and deemed the *Albert Offset* the best approach [11]. In contrast, Grill *et al*. (2016) described this approach as the worst-performing one in their study [12]. These differences may relate to the *Albert Offset* approach’s strong assumption that the covariate distribution is the same in the training dataset as in the population for which the probabilities are adjusted [10, 11], and as we showed in sFig. 5, the *Albert Offset* approach can perform poorly for datasets where the covariate distribution is different. When recalibrating models for different settings, the likelihood ratio assumption could become harder to justify depending on the specific setting, and particular caution would be required in populations where the clinical characteristics of the patients differ substantially from the case-control dataset used for original model development. Establishing whether the similarities between covariate distributions are sufficient for pre-assessing the performance of the *Albert Offset* method is of interest for future research.

When only a population-representative dataset is available, the *Re-estimation* approach would be necessary. The *Re-estimation* approach calibrates well in the general population dataset for both scenarios but demonstrates high uncertainty surrounding the model predictions. In contrast, Grill *et al*. (2016) describe the *Re-estimation* approach as having equivalent performance to the *Albert Offset*, with both performing worse than all other approaches [12]. The high uncertainty in our analysis can be attributed to the fact that there are only 7 positive MODY cases in the general population dataset (prevalence of 0.6%) and that the distribution of predicted probabilities is skewed towards zero. This highlights the problem with fitting models for rare diseases to population data [31], where the low prevalence means very large sample sizes would be required to reduce the uncertainty around the predictions, and it would be important to assess the adequacy of the sample size prior to model fitting [32]. When both training and population-representative datasets are available (as in our study), we showed that the *Recalibration* approach demonstrates good calibration in the population-representative dataset for both scenarios. This approach combines the information captured from the training data with information from the calibration dataset [7, 33], producing relatively low uncertainty in the model predictions compared to other approaches explored in this paper.

We also explored a scenario where additional biomarker testing was available but performed only on a limited subset of patients. Screening using biomarkers is common in clinical practice and often used in rare diseases where universal testing is not cost-effective or could be invasive (e.g. screening for chromosomal defects in pregnancy [34]). We developed a Bayesian hierarchical mixture model to follow the referral process involved in MODY testing and, therefore, utilise the additional biomarker tests for further refinement in the prediction of MODY probabilities. As a Bayesian model, we can incorporate additional information from other studies into the prior distributions for certain parameters, something previously explored by Boonstra *et al*. [15] in a different setting where additional information is only present for a subset of individuals. This approach has a further advantage in that predictions can still be made for patients with missing additional biomarkers, which are modelled using patient characteristics. This is important for our setting in which instead of ignoring the biomarker results altogether, the model has used this information to improve predictions so that even when biomarker information is missing, the MODY probabilities are a weighted sum across the latent biomarker test results, where the weights are informed by a model relating potential biomarker test outcomes conditional on a set of clinical features. We also combined the mixture model with the *Re-estimation* and the *Recalibration* approaches for just *C*^+^ ∩ *A*^−^ individuals. Both approaches showed uncertainty levels in the probability predictions consistent with the previously observed uncertainty estimates. Furthermore, both approaches were tested for stability using bootstrapped versions of the population-representative dataset [35], demonstrating that the *Recalibration mixture* approach was more stable with the predicted probabilities of MODY than the *Re-estimation* mixture approach.

Other approaches for recalibration have been considered in previous work. Chan *et al*. (2008) [11] compared three methods to update pre-test probability with information on a new test: the Albert (*Albert Offset* in our study) [10], Spiegelhalter and Knill-Jones (SKJ) [36] and Knottnerus [37] approaches. The SKJ represents an alternative to the Albert Offset approach, with similar performance in their paper. The Knottnerus approach was more suited to cases with sequential biomarker testing, which was not appropriate for our work since we did not have data on some combinations of tests, instead we grouped biomarkers into a composite measure *T*. The Knottnerus approach could be compared to the mixture model approaches (allowing for non-independence between both biomarkers), and examining these approaches when considering sequential testing of more than one biomarker could be considered in future work. Grill et al. (2016) compared several methods for incorporating new information into existing risk prediction models: logistic-new (equivalent to *Re-estimation*), LR-joint, LR-offset (*Albert Offset*), and LR-shrink (equivalent to SKJ from reference [11, 36]). In contrast to our study, their original models were built in population data, and the new datasets with additional features were either cohort or case-control data [12]. In the context of rare diseases, case-control data is likely to provide the best dataset for initial model development since this gives the most power for estimating model parameters, and the population-representative model can then borrow information from the case-control model. In cases where the additional data are only available in the case-control setting, and original models were built on population data, the joint model approach (*Recalibration*) could be adaptable to this scenario.

The model we recommend for the available MODY data is the *Recalibration mixture* approach. A major strength of this procedure is that it allows predictions for patients with missing biomarker testing, and this weighted prediction of MODY probability can be used to inform whether a patient should be referred for further testing [38]. This model allows for the incorporation of strong prior information regarding the probability of having MODY for *C*^−^ ∪ *A*^+^ individuals, propagates uncertainties regarding the missing data in the UNITED study, and borrows weight from the case-control model through the recalibration procedure [7], thus improving the stability of predictions [35]. This model provides sensible predictions for the probability of MODY for patients with/without additional testing for C-peptide and antibodies. Patients with missing MODY testing (i.e. *C*^+^ ∪ *A*^−^) could have been set as a negative result test for all approaches due to the strong clinical knowledge of these tests being consistent with a T1D diagnosis. However, this may not be the case for other settings, where patients with missing outcomes could be believed to have a higher probability of the outcome, and therefore, assuming that the outcome is negative may be less justifiable.

There are some limitations to the *Recalibration Mixture* approach. We currently use biomarker tests as binary (positive/negative) results; in practice, biomarkers may be on a continuous scale. As such, the model could be adapted to include the biomarker results as additional covariates, which could be numerically integrated out if predicting to an individual that was missing this information in practice [38]. We are also limited by the small sample sizes in rare diseases [39], and even with our final model utilising two datasets, model predictions still have some uncertainty. However, we still saw good separation between MODY and T1D, and even accounting for the uncertainty, probability thresholds could be defined that rule out clear non-MODY cases and can be used to determine positive test rates at different probabilities in practice. These thresholds would balance the amount of testing to be carried out against the potential for missing genuine MODY cases, depending on how conservative the choice of threshold is. The model has yet to be validated in a hold-out dataset, but the stability plots using bootstrapped datasets provide some insight into the stability of model predictions [35]. Although the 95% credible interval of bootstrapped probabilities is relatively wide at higher values, the 50% credible interval is narrow for all probabilities around the equal line.

This paper provides a comparison of several recalibration approaches. The development of our recalibration approach uses established methodologies, and we have shown how it could apply to identifying patients with a high probability of MODY to allow more targeted diagnostic testing, but these ideas could be applied to other diseases. In practice, other settings could benefit from a similar Bayesian hierarchical model structure where informative biomarkers or additional testing information are available but only in a subset of patients due to its invasive nature or high cost of testing. With this structure, the model can be used to consider whether additional testing should be carried out when the individual already has a low probability (not on the cusp of referral), something explored previously in treatment selection for Type 2 diabetes [38]. Furthermore, this modelling structure could be particularly useful in other rare diseases with low sample sizes since it borrows weight from multiple datasets through recalibration, improving predictions.

## 5. Conclusion

We have compared several approaches to developing prediction models for rare diseases. We found the *Recalibration mixture* model approach to be the best approach, combining case-control and population-representative data sources. This approach allows the incorporation of additional data on biomarkers and appropriate prior probabilities.

## Data Availability

Data are available through application to the Genetic Beta Cell Research Bank https://www.diabetesgenes.org/current-research/genetic-beta-cell-research-bank/ and the Peninsula Research Bank https://exetercrfnihr.org/about/exeter-10000-prb/. Example R code for fitting the approaches used in this study is available on GitHub within the repository "Exeter-Diabetes/Pedro-MODY_recal_approaches".

https://github.com/Exeter-Diabetes/Pedro-MODY_recal_approaches

## 6. Ethics approval and consent to participate

For the UNITED study, ethics approval was granted by the NRES Committee South West–Central Bristol (REC no. 10/H0106/03). For the case-control data, this study was approved by the Genetic Beta Cell Research Bank, Exeter, UK (ethical approval was provided by the North Wales Research Ethics Committee, UK (IRAS project ID 231760)) and the Peninsula Research Bank (Devon & Torbay Research Ethics Committee, ref: 2002/7/118).

## 7. Consent for publication

Not applicable.

## 11. Authors’ contributions

PC, BMS and TJM (McKinley) conceived and designed the study. PC, BMS and TJM (McKinley) analysed the data and developed the code. KAP and TJM (McDonald) provided the cohort data partly used to estimate the prior distribution of a positive MODY test result, into those with C-peptide negative or autoantibody positive test results. ATH and ERP led the UNITED population study. All authors contributed to the writing of the article, provided support for the analysis and interpretation of results, critically revised the article, and approved the final article.

## 12. Acknowledgements

For the purpose of open access, the author has applied a CC BY public copyright license to any Author Accepted Manuscript version arising from this submission.

