## Supplementary Material for "Comparison of Bayesian approaches for developing prediction models in rare disease: application to the identification of patients with Maturity-Onset Diabetes of the Young"

### SM Methods:

#### Notation

This section contains a glossary for all mathematical symbols used throughout the manuscript.

$C$ – C-peptide test, with superscript corresponding to the test result (^+^ for positive, ^-^ for negative)

$A$– Autoantibody test, with superscript corresponding to the test result (^+^ for positive, ^-^ for negative)

$C^{-}\cup A^{+}$ – combination of C-peptide negative or autoantibody positive test results

$C^{+}\cap A^{-}$ – combination of C-peptide positive and autoantibody negative test results

$T_{i}^{U}$ – combination of C-peptide and autoantibody test results

$M$ – MODY test, with superscript corresponding to the test result (^+^ for positive, ^-^ for negative)

$R$ – hypothetical test, with superscript corresponding to the test result (^+^ for positive, ^-^ for negative)

$M_{j}^{C}$ – MODY test, with superscript $C$ corresponding to the case-control dataset and the subscript $j$ representing the patient of case-control dataset

$M_{i}^{U}$ – MODY test, with superscript $U$ corresponding to the UNITED dataset and the subscript $i$ representing the patient of UNITED dataset

$X_{jv}^{C}$ – a set of $p$ covariates ($v=1,\ldots,p)$ for individual $j$ of the case-control dataset

$X_{iv}^{U}$ – a set of $p$ covariates $\left( v=1,\ldots,p \right)$ for individual $i$ of the UNITED dataset

$X^{*U}$ – a set of covariates used to model the variable $T_{i}^{U}$

$\beta^{C}$ – model parameters for a model fitted with case-control dataset

$\beta^{U}$ – model parameters for a model fitted with UNITED dataset

#### Prior elicitation

A major advantage of the *mixture model* approach is formally building in appropriate low prior probabilities for the individuals with C-peptide negative or antibody positive test results rather than just considering all as $\text{MOD}\text{Y}^{-}$. The prior probability of $M^{+}|C^{-}\cup A^{+}$ can be modelled as:

$$P\left( M^{+} | C^{-}\cup A^{+} \right)\propto P\left( C^{-}\cup A^{+} | M^{+} \right)P\left( M^{+} \right), (1)$$

from a straightforward application of Bayes’ Theorem. We can estimate $P\left( C^{-}\cup A^{+} | M^{+} \right)$ using data from the “One tube study”, which tested C-peptide and islet autoantibodies in sequential referrals to the UK MODY diagnostic genetic testing service within the Exeter Genomics Laboratory at the Royal Devon and Exeter National Health Service (NHS) Foundation Trust, which provides monogenic diabetes testing for England, Wales, and Northern Ireland. In this cohort, in 282 individuals who tested positive for MODY, only 6 were $C^{-}\cup A^{+}$(personal communication T McDonald). Under a flat U(0, 1) prior distribution for $P\left( C^{-}\cup A^{+} | M^{+} \right)$, we can fit a binomial model to these data giving a posterior distribution:

$$P\left( C^{-}\cup A^{+} | M^{+} \right) \sim\text{Beta}\left( \alpha=7, \beta=277 \right) (2)$$

Furthermore, we can estimate $P(M^{+})$ (the prevalence of MODY in young-onset insulin-treated patients) from an independent UK population screening study [1], which found 2 individuals with MODY out of 237 individuals diagnosed with T1D. Again, under a flat U(0, 1) prior distribution for $P\left( M^{+} \right)$, we can fit a binomial model to these data giving a posterior distribution:

$$P\left( M^{+} \right) \sim\text{Beta}\left( \alpha=3, \beta=246 \right) (3)$$

These two distributions in (2) and (3) can then be used to generate an estimate for (1) using Monte Carlo simulation. Here we take independent random samples from (2) and (3), and thus derive random samples for $P\left( M^{+} | C^{-}\cup A^{+} \right)$ by taking their product. We then fit a Beta distribution to these random samples using maximum likelihood, and we can use this fitted beta distribution as a tractable prior distribution for (1) in our MODY model, giving:

$$P\left( M^{+} | C^{-}\cup A^{+} \right) \sim\text{Beta}\left( \alpha=2.2, \beta=7361.3 \right) (4)$$

We note that $E\left[ P\left( M^{+} | C^{-}\cup A^{+} \right) \right]$ is very small here and is in line with the choice not to genetically test for MODY for $C^{-}\cup A^{+}$ individuals during the UNITED study, however using this Beta prior is more conservative than assuming a prior probability of exactly zero. Furthermore, the distributions in (2) and (3) were generated using flat uniform priors and so (4) should be conservative in the sense of having the largest variance compared to other choices of prior distributions. A plot of the histogram of the random samples versus the fitted Beta distribution is shown in sFig. 6.

#### Stability analysis

We compared model stability for the *Re-estimation* and *Recalibration* mixture approaches to investigate the impact of low-case numbers. The study investigated the instability of the model by fitting the same model to 1,000 bootstrapped datasets [2]. The instability is shown by deriving a prediction instability plot of bootstrap model predictions (y-axis) versus original model predictions (x-axis). Stability is demonstrated when most values land close to the diagonal line (y=x).

### References

| [1] | G. Thanabalasingham, A. Pal, M. P. Selwood, C. Dudley, K. Fisher, P. J. Bingley, S. Ellard, A. J. Farmer, M. I. McCarthy and K. R. Owen, “Systematic assessment of etiology in adults with a clinical diagnosis of young-onset type 2 diabetes is a successful strategy for identifying maturity-onset diabetes of the young,” *Diabetes Care,* vol. 35, no. 6, pp. 1206-1212, 2012. |
| --- | --- |
| [2] | R. D. Riley and G. S. Collins, “Stability of clinical prediction models developed using statistical or machine learning methods,” *Biometrical Journal,* p. 2200302, 2023. |

### SM Results:

**sTable 1: Model parameter estimates of the T model for *Re-estimation* and *Recalibration mixture* approaches for scenario c).** Scenario c): analyse all patients (n=1,171). The numbers in the brackets correspond to the 95% credible intervals.

| Model parameters | Re-estimation mixture | Recalibration mixture |
| --- | --- | --- |
| Intercept | 2.82 (0.49;5.20) | 2.81 (0.35;5.25) |
| BMI (kg/m^2^) | 0.07 (-0.05;0.19) | 0.07 (-0.06;0.20) |
| Spline(BMI) | -0.13 (-0.25;0.01) | -0.13 (-0.25;0.00) |
| Age at diagnosis (years) | -0.34 (-0.50;-0.20) | -0.33 (-0.49;-0.19) |
| Spline(Age at diagnosis) | 0.23 (0.08;0.38) | 0.22 (0.08;0.37) |
| Age at recruitment (years) | 0.10 (0.03;0.17) | 0.10 (0.02;0.16) |
| Spline(Age at recruitment) | -0.05 (-0.13;0.02) | -0.05 (-0.13;0.02) |
| At least one parent affected with diabetes | -1.03 (-1.50;-0.54) | -1.03 (-1.50;-0.54) |

**sFig. 1: Characteristics of patients in the case-control dataset and the UNITED dataset. The diagonal contains density plots (continuous) and bar plots (categorical) comparing a variable between datasets.** The lower triangle contains density plots (continuous versus categorical), 2-dimensional density plots (continuous versus continuous) and bar plots (categorical vs categorical) comparing variables between datasets. The case-control dataset is represented with red, and the UNITED dataset is represented with blue.

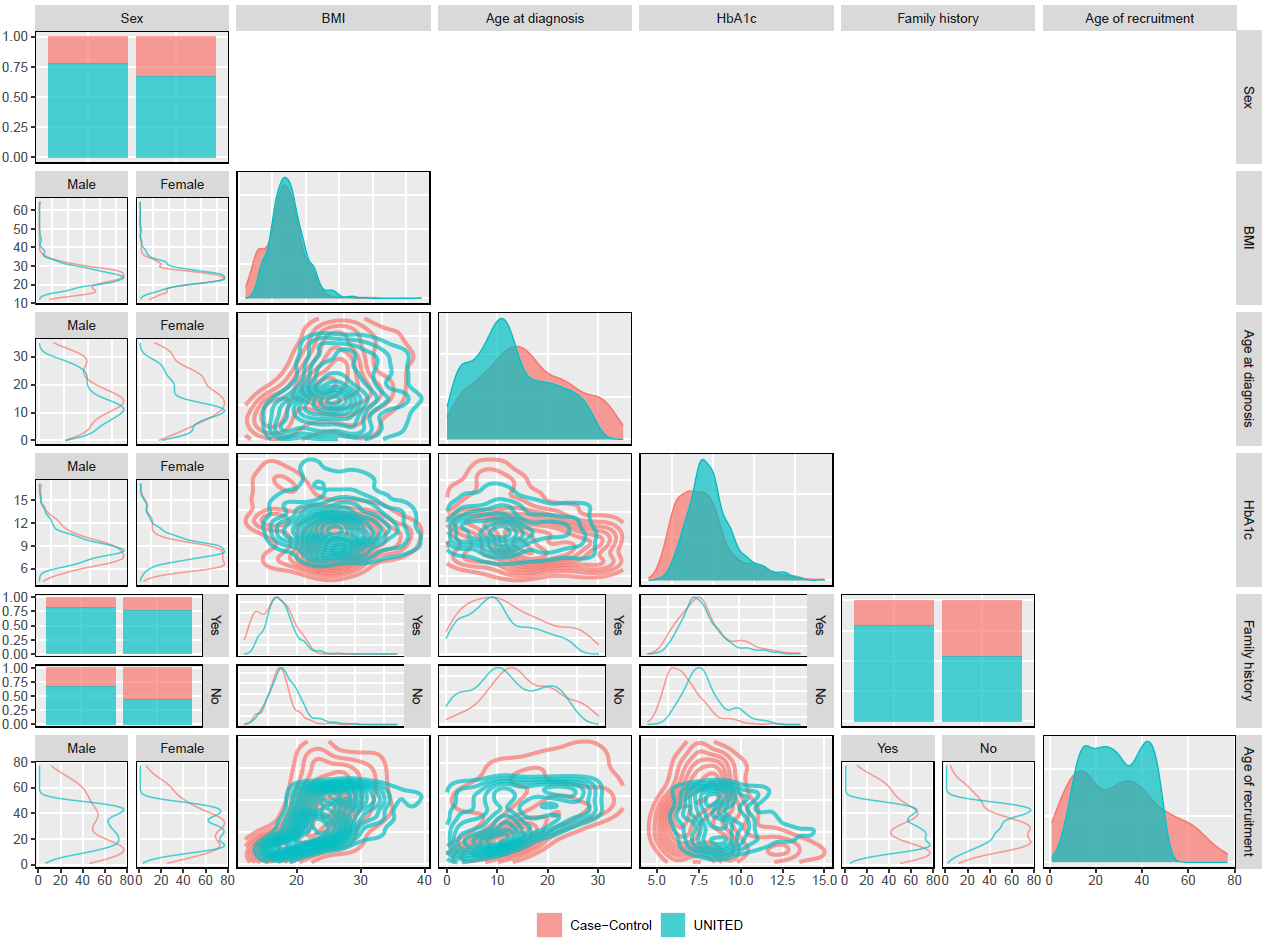

**sFig. 2: Trace plots of regression parameters for 4 chains with 500,000 iterations (300,000 iterations of burn in removed) in scenario a).** A) regression parameters for the *Original* and *Albert Offset* approaches. B) regression parameters for the *Re-estimation* approach. C) regression parameters for the *Recalibration* approach.

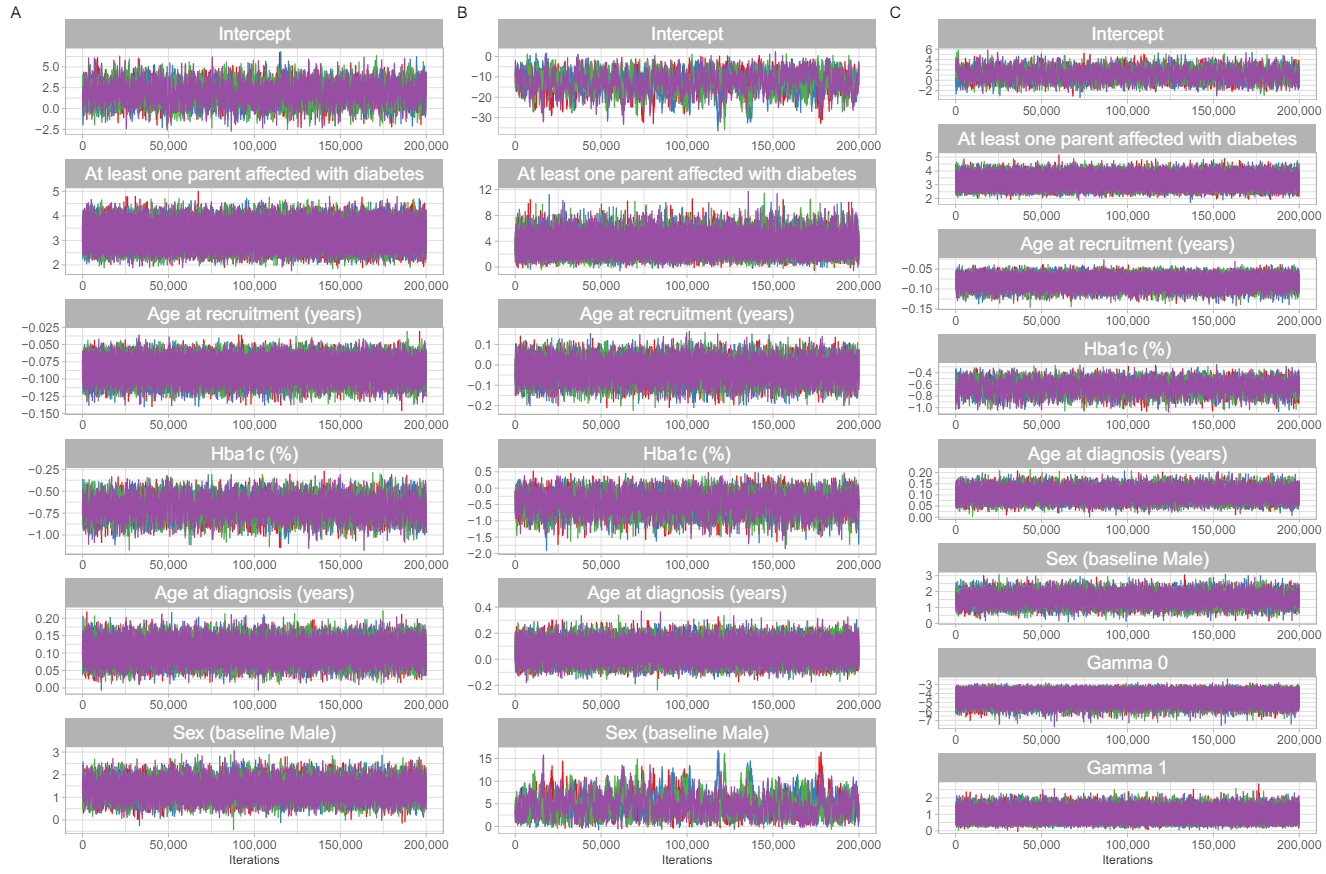

**sFig. 3: Trace plots of regression parameters for 4 chains with 500,000 iterations (300,000 iterations of burn in removed) in scenario b).** A) regression parameters for the *Original* and *Albert Offset* approaches. B) regression parameters for the *Re-estimation* approach. C) regression parameters for the *Recalibration* approach.

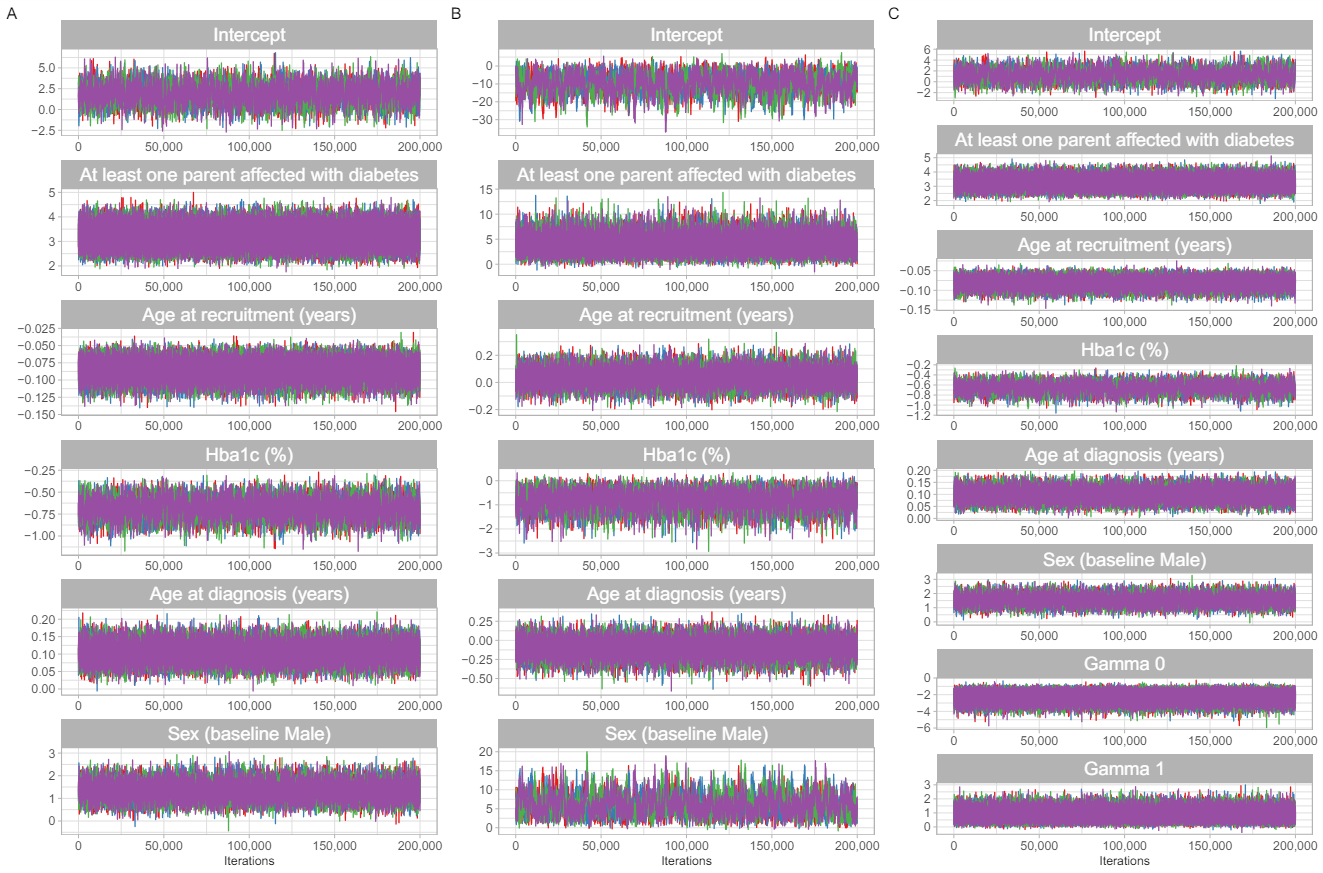

**sFig. 4: Trace plots of regression parameters for 4 chains with 500,000 iterations (300,000 iterations of burn in removed) in scenario c).** A) regression parameters for the *Re-estimation Mixture* approach. B) regression parameters for the *Recalibration Mixture* approach.

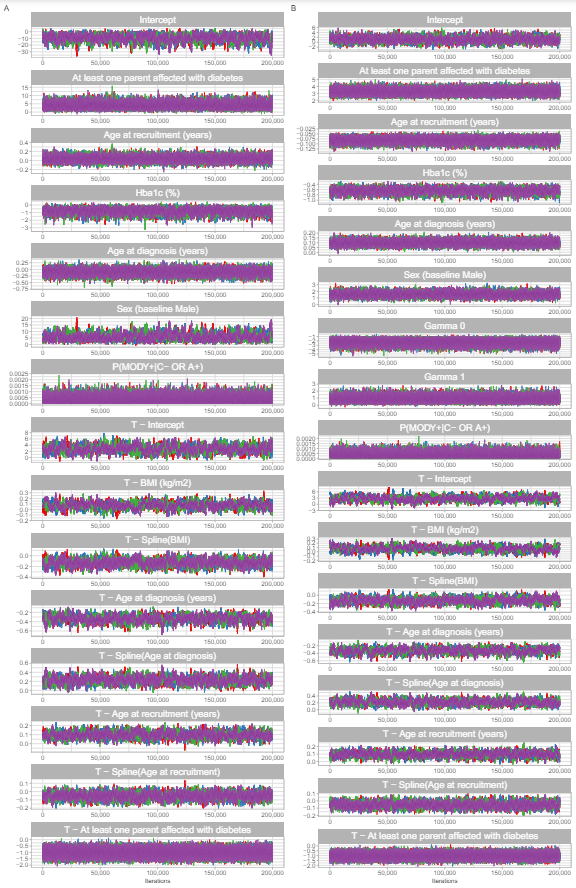

**sFig. 5: Calibration of Albert Offset and Recalibration methods in the same and different likelihood ratios.**

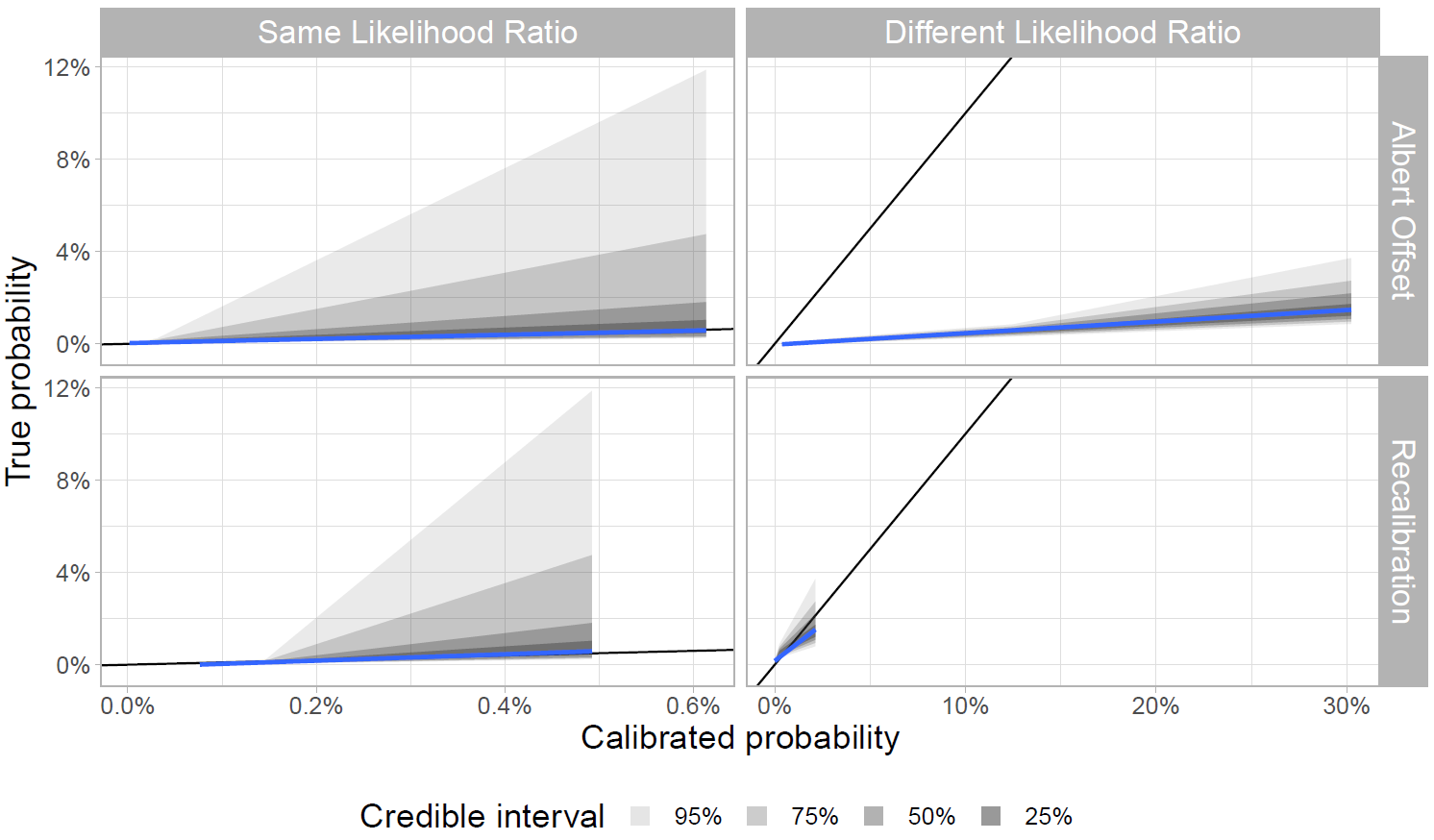

**sFig. 6: Prior distribution placed on** $\mathbf{C}^{\mathbf{-}}\boldsymbol{\cup}\mathbf{A}^{\mathbf{+}}$ **patients in the mixture model approach c).**

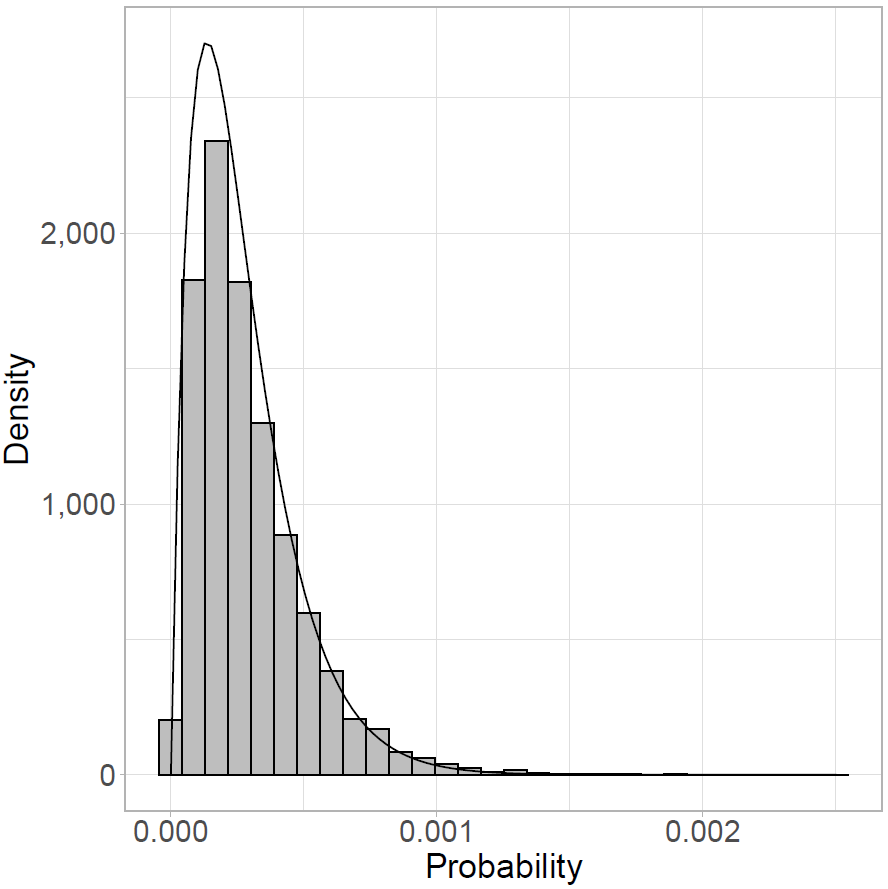
